# Statistical Analysis Plan for the INTIMET study (Insulin Resistance in Type 1 Diabetes Managed with Metformin)

**DOI:** 10.1101/2023.11.20.23298602

**Authors:** Jennifer R Snaith, Nick Olsen, Jerry R Greenfield

## Abstract

This document provides the full statistical analysis plan (SAP) for the INTIMET study (Insulin Resistance in Type 1 Diabetes Managed with Metformin), a randomised double-blinded placebo-controlled trial, designed to evaluate the effect of metformin on insulin resistance and cardiometabolic health in type 1 diabetes. This trial was prospectively registered within the Australian and New Zealand Clinical Trials Registry (ACTRN12619001440112). The study protocol has previously been published (Snaith JR et al, Diabetic Medicine 2021).

## 1. SECTION 1: ADMINISTRATIVE INFORMATION

### 1.1. Title and trial registration

The INTIMET study (<u>In</u>sulin Resistance in <u>Type 1</u> Diabetes Managed with <u>Met</u>formin) is a parallel group, superiority, randomised double-blind placebo-controlled trial examining whether metformin is superior to placebo to reduce insulin resistance and improve cardiometabolic parameters in type 1 diabetes. Predictors of response to metformin will also be assessed. INTIMET includes a cross-sectional study of both type 1 diabetes with age-BMI and gender-matched controls without diabetes to enable a detailed study of the phenotype of insulin resistance in type 1 diabetes.

Trial registration:

Australian New Zealand Clinical Trials Registry (ANZCTR) ACTRN12619001440112

First registered: 17/10/2019

Universal trial number: U1111-1238-8090

### 1.2. SAP version

SAP version 2.0, Dated 24/1/23. This SAP version follows guidelines for SAP reporting (Gamble et al. JAMA 2017).^1^

### 1.3. Protocol version

This SAP is based on the original protocol approved by the St Vincent’s Hospital Human Health Research Ethics Committee, approved May 2019 (2019/ETH00379, up to Version 8 approved 16^th^ Dec 2021) and the published study protocol (https://doi.org/10.1111/dme.14564).^2^

### 1.4. SAP REVISION HISTORY

SAP revision history:

- Original version: published study protocol, submitted 14/12/2020, accepted 24/3/2021
- Latest study protocol: version 8 23/11/2021
- Original main statistician: Zhixin Liu
- Change in main statistician: Nick Olsen (2/12/2022)

Justification of revisions:

| Relevant SAP section (v2.0) | Change made | Reason |
| --- | --- | --- |
| 1.5 – roles and responsibilities | Change in study statistician | Change in UNSW staffing |
| 4 – statistical principles | Increased detail on specifics of statistical plan | Original SAP did not provide full level of detail. |
| 6.1 – outcome definitions | Revising low-dose EGP as the key primary endpoint | Reduce multiplicity with both basal Ra and low-dose EGP, and the measure of low-dose EGP is more robust than basal Ra in type 1 diabetes. |
|  | Change from ANCOVA to generalised linear mixed modelling as the primary analysis method. | Advice following change in statistician that GLM provides precise parameter estimates than ANCOVA for the primary outcome assessment. |

### 1.5. Roles and responsibilities

Names, affiliations, and roles of SAP contributors:

**Prof Jerry Greenfield**, MBBS (Hons), PhD, FRACP

Garvan Institute of Medical Research, University of New South Wales, Sydney, Australia, St Vincent’s Hospital Sydney

Responsibility: Chief Investigator

**Dr Jennifer Snaith**, MBBS(Hons), BMedSc, FRACP

Garvan Institute of Medical Research, University of New South Wales, Sydney, Australia, St Vincent’s Hospital Sydney

Responsibility: Preparation of SAP, study coordinator, performed all studies and data collection

**Dr Nick Olsen**, BSci (Hons), PhD

Mark Wainwright Analytical Centre, University of New South Wales, Sydney, Australia

Responsibility: Lead Statistician

## 2. SECTION 2: INTRODUCTION

### 2.1. Background and rationale

Type 1 diabetes affects 120 000 individuals in Australia and the incidence is rising.^3^ It is a condition typified by autoimmune destruction of pancreatic beta cells leading to insulin deficiency necessitating insulin replacement. People with type 1 diabetes have an increased cardiovascular (CV) risk and excess mortality relative to general population. However, this increased risk is not explained by hyperglycaemia alone.

Insulin resistance is under recognised in type 1 diabetes and is relevant to altered metabolic pathways in fat, liver, muscle and other tissues. The Garvan laboratory has previously established that type 1 diabetes individuals are more insulin resistant than their non-diabetic peers. This was determined using insulin clamp studies, the gold standard measure of insulin resistance.^4^ However, there remain key unanswered questions with regards to insulin resistance in type 1 diabetes, including its cause, its consequences and its phenotype including whether it arises from liver, muscle or both. Heterogeneity in the degree and organ source of insulin resistance may explain diversity in metabolic phenotypes in type 1 diabetes, where some individuals are predisposed to metabolic syndrome with weight gain, increased insulin requirements, adverse lipid and blood pressure profiles and increased cardiovascular risk. This is also strongly suggested by the finding that type 1 diabetes patients with greater clamp-derived insulin resistance associate with greater degrees of coronary artery calcification.^5^ Heterogeneity in insulin resistance may also explain findings of variable responses to metformin in type 1 diabetes.

Metformin is an anti-hyperglycaemic agent that improves glycaemia in type 2 diabetes by lowering hepatic glucose output, and increasing peripheral glucose uptake in muscle.^6^ It is thought to be beneficial to CV outcomes in type 2 diabetes.^5^ Metformin has potential for repurpose in type 1 diabetes. Disappointingly, metformin studies in type 1 diabetes have not demonstrated consistent results, and at most have shown only minor improvements in weight, cholesterol, glycaemia and no clear reduction in CV risk.^7^ Although insulin replacement has been the mainstay of treatment in type 1 diabetes, there may be rationale for adjunctive agents to address cardiometabolic risk and may reduce insulin resistance.

#### Study rationale

The role and pathogenesis of insulin resistance in type 1 diabetes is not well understood. It has been established that type 1 diabetes individuals are more insulin resistant than normal individuals, but the organ source of insulin resistance (muscle vs liver vs both) is not well characterised. Furthermore, the phenotype of insulin resistance, and whether liver vs muscle insulin resistance has consequences for lipid and blood pressure profiles, weight, or cardiovascular risk requires further study.

Our trial is the first study to examine organ specific patterns of insulin resistance in adults with type 1 diabetes and is the first to study heterogeneity in insulin resistance and response to metformin.

This study has the potential to change treatment paradigms with an increased focus on treatment of insulin resistance in type 1 diabetes, and to define whether tissue specific insulin resistance is a candidate target for adjunctive therapy.

### 2.2. Aims and objectives

#### Hypotheses

1. Type 1 diabetes is associated with heterogeneous patterns of insulin resistance in muscle and liver (compared to matched controls).
2. In type 1 diabetes, insulin resistance aligns with worse metabolic and cardiovascular profiles (higher blood pressure, adverse lipid profile, visceral and hepatic steatosis).
3. Liver, but not muscle, insulin resistance is a determinant of metformin’s effect in type 1 diabetes.
4. Treatment with metformin will improve insulin sensitivity in T1D in participants displaying hepatic insulin resistance.

#### The overall aims of this project are to

1. Characterise patterns of insulin resistance in muscle and liver in type 1 diabetes and determine the impact on metabolic phenotype.
2. Investigate whether metformin improves insulin resistance in type 1 diabetes and to determine the predictors of response to metformin in type 1 diabetes.

#### Specific aims include: -- check these are all answered

1. To determine whether muscle and/or liver insulin resistance is present universally or selectively in people with type 1 diabetes.
2. To determine whether site of insulin resistance (liver vs muscle vs both) in type 1 diabetes cluster with adverse metabolic and cardiovascular risk factors, and bone parameters (density and turnover)
3. To identify non-invasive clinically available markers that identify individuals with type 1 diabetes with increased insulin resistance.
4. To determine whether metformin improves insulin resistance at muscle and/or liver (primary outcome) and other metabolic and vascular measures (secondary outcomes – see below) in type 1 diabetes.
5. To determine the relationship between organ specific insulin resistance and response to metformin in type 1 diabetes.
6. To determine which individuals with type 1 diabetes most improve their clamp-derived measures of insulin resistance following treatment with metformin.
7. To create predictive algorithms using non-invasive measures that identify individuals with type 1 diabetes with increased insulin resistance, and individuals likely to respond to metformin.

## 3. SECTION 3: STUDY METHODS

### 3.1. Trial design

The INTIMET study has a 2-part design:

#### Part 1

Cross-sectional study of 40 adults with type 1 diabetes and 20 adults as a non-diabetes comparator group (2:1 ratio) to extensively characterise the phenotype of insulin resistance in adults with type 1 diabetes.

#### Part 2 (main study)

Double blinded randomised placebo-controlled trial of 6 months of metformin therapy (1.5g XR daily) in adults with type 1 diabetes (1:1 randomisation).

The study design has been developed in accordance with:

- SPIRIT statement for interventional trials: http://www.spirit-statement.org/
- CONSORT guidelines for randomised trials: http://www.consort-statement.org/
- STROBE criteria for observational studies: https://www.strobe-statement.org

### 3.2. Randomisation

Randomisation of metformin to placebo will be 1:1 using minimisation (Minitable program) modified from Saghaei et al ^8^. Factors will be weighted for importance and included BMI (40% weighting), HbA1c (20% weighting), gender (20% weighting), age (20% weighting). The factors will be provided to a clinician independent to the study who will then perform the randomisation and communicate the allocation directly to the St Vincent’s Hospital clinical trials pharmacy.

Research staff (including those involved with recruitment and outcome assessors), participants, care providers and data analysts will remain blinded to the treatment allocation. The dispensing pharmacist will not have contact with study participants.

The central study coordinator will code the 2 treatment arms as Group A and Group B (determined by coin toss) before providing the data to the statistician.

### 3.3. Sample size

A sample size of 40 participants (20 per treatment arm) was determined to provide 80% power at a two-sided significance level of 0.05 to detect a mean difference of 0.3 mg/kg/min with a pooled standard deviation of 0.3 mg/kg/min (effect size of 1) in the primary outcome measure of Ra (a measure of endogenous glucose production; EGP) at week 26, assuming a 15% attrition rate. This effect size estimation was taken from Cree-Green et al, a study of the effect on EGP in an adolescent population with type 1 diabetes treated with metformin for 3 months.^9^

Note: this updated statistical analysis plan included refinement in the primary outcome measure definition, from both basal Ra and low-dose EGP to low-dose EGP alone as a single primary outcome measure. The consolidated primary outcome was adopted after the protocol was published and registered in ANZCTR but before the final data was analysed. The study sample size was not updated.

### 3.4. Framework

Primary and secondary outcomes will be assessed using a superiority framework, expecting that participants receiving metformin will improve more than those receiving placebo.

### 3.5. Statistical interim analysis and stopping guidance

No stopping rule is defined. Recruitment will continue until the target of n=40 participants with type 1 diabetes is reached. This is to ensure a power of 80% anticipating a potential 15% loss to follow-up.

### 3.6. Timing of final analysis

The first mid-trial follow-up visit is at 13 weeks, with the main follow up visits at 26 weeks after commencing treatment. All primary outcomes will be analysed by an independent statistician who is blinded to the allocation (N.Olsen). Secondary analyses will either be analysed by the statistician or by other study investigators under the supervision of the blinded statistician.

Data from all time points (baseline, 13 weeks, 26 weeks) will be included in the analysis. Analysis will occur after the final volunteer completes their final study visit and all tracer clamp data (required for primary outcome data) is processed and available for interpretation.

### 3.7. Timing of outcome assessments

Timing of outcome assessments per Table 3 of published protocol paper.^2^

## 4. STATISTICAL PRINCIPLES

### 4.1. Confidence intervals and p-values

All assessments of between group effects will use two-sided tests with a 5% significance level, p=0.05. Confidence intervals will be presented as 95% and two-sided.

Regarding secondary outcome analysis, the number of statistically significant analysis that we expect from chance alone based on the number of subgroup analysis will be acknowledged in data interpretation. The primary outcome is well-defined (change in low-dose EGP from baseline to 26 weeks).

### 4.2 Adherence and protocol deviations

Adherence to intervention will be assessed by tablet count upon final tablet return. Poor compliance will be defined as return of >25% of the target tablets for the max tolerated dose of that individual.

A protocol deviation will be defined as >15% loss to follow up or less than 40 volunteers with type 1 diabetes undergoing randomisation. Any protocol deviations will be reported in the final report.

### 4.3 Analysis populations

Analysis of all randomised subjects will be conducted using an intention-to-treat approach (ITT) according to the treatment they were allocated to receive. This will be the primary population for the analysis.

If after assessing the nature of missing data, if we deem that data is missing completely at random (MCAR), then complete case analysis will be used for the primary analysis. Subjects with missing data on any variables used in models will be excluded from that analysis. If missing at random, then we will perform multiple imputation.

Subgroups: there are numerous subgroups of interest as subgroup analysis formed part of the main study hypothesis and primary analysis.

Baseline data will be divided into 2 subgroups according to the presence or absence of type 1 diabetes.

The population with type 1 diabetes will be divided into insulin resistant vs insulin sensitive by tissue site

a. Liver insulin resistant: defined as EGP above the median LD-EGP of group with type 1 diabetes
b. Muscle insulin resistant: defined as GIR below the median GIR of the group with type 1 diabetes.

Select analysis will be divided into subgroups such as by gender subgroups, BMI categories or the presence/absence of medications (eg. statins). Subgroup analyses will be reported per guidelines and the impact of multiple subgroup analyses will be considered and reported.^10^

Where SHBG is included in a model, women taking hormone-based contraception will be excluded.

## 5. SECTION 5: TRIAL POPULATION

### 5.1. Screening data

The time frame for recruitment (start and end date), first and last participant enrolment and data collection will be reported. The total number of subjects screened for eligibility will be declared in the CONSORT diagram.

### 5.2. Eligibility

#### Summary of eligibility criteria

##### Inclusion

- Age range: 20-55 years (and pre-menopausal if female)
- Type 1 diabetes:
  1. Disease duration > 10 years (ie. chronic exogenous insulin exposure)
  2. Insulin deficiency (fasting c-peptide < 0.3nmol/L)
  3. HbA1c less than or equal to 9.5%

##### Exclusion

1. Current smoking
2. Current or planned prescription of medications that affect glucose metabolism (glucocorticoids, antipsychotics, immunosuppressants).
3. Exposure to metformin within the last 30 days
4. Alcohol intake > 20g/day in women or > 40g/day in men
5. Weight change > 5% in last 3 months or history of bariatric surgery
6. Pregnancy, breastfeeding, or childbearing potential not willing to avoid pregnancy during the study.
7. Known major organ dysfunction (eGFR < 60, liver disease transaminases > 3 times the upper limit of normal, cardiac event within the last 6 months, current cancer or uncontrolled thyroid dysfunction).
8. Diabetic ketoacidosis or severe hypoglycaemia (hypoglycaemia requiring third-party assistance) in the last 6 months.
9. A history of a psychological illness or condition that would interfere with the patient’s ability to understand the requirements of the study.

### 5.3. Recruitment and withdrawal/ follow up

The following information will be included in the CONSORT flow diagram:

- all volunteers assessed for eligibility
- all volunteers meeting exclusion criteria, and the reason for exclusion
- all volunteers eligible for inclusion
- all volunteers not consenting (and reasons)
- all patients randomised
- all patients that undergo assessment at 3 and 6 months
- withdrawals with reasons and timing and treatment arms
- patients included in the ITT, per protocol and as treated analyses in both treatment arms

### 5.4. Baseline patient characteristics

List of baseline characteristics to be summarised:

Age (years)

Sex (% male)

HbA1c (%, mmol/mol)

BMI (kg/m^2^)

SBP (mmHg)

DBP (mmHg)

Diabetes duration (years)

Ethnicity (% Caucasian)

Family history ischaemic heart disease (n,%)

Family history type 2 diabetes (n,%)

Waist to hip ratio

Insulin delivery method

Pump (n, %)

MDI (n, %)

Total daily insulin dose (U/Day)

Time in range (%)

Time in hyperglycaemia (%)

Time in hypoglycaemia (%)

Glycaemic variability (CV)

Retinopathy grading (n,%)

Maculopathy grading (n,%)

Augmentation index (AIx)

CAP (dB/m)

MRI liver fat (%)

MRI muscle fat infiltration (%)

MRI abdominal subcutaneous adipose issue volume (L)

DXA visceral adipose tissue (g)

DXA total fat mass (kg)

DXA total fat free mass (kg)

Total cholesterol (mmol/L)

HDL (mmol/L)

LDL (mmol/L)

Triglycerides (mmol/L)

Bilirubin (μmol/L)

Total protein (g/L)

Albumin (g/L)

Total globulin (g/L)

ALT (U/L)

AST (U/L)

GGT (U/L)

ALP (U/L)

Uric acid (mmol/L)

IL-6 (pg/mL)

sICAM-1 (ng/mL)

SE-selectin (ng/mL)

Adiponectin (μg/mL)

IGF1 (nmol/L)

SHBG (nmol/L)

GDF-15 (pg/mL)

These will be presented as type 1 diabetes vs control for the observational study, and as metformin vs placebo for the main RCT.

The number of available measurements of the baseline characteristics will be reported.

## 6. ANALYSIS

### 6.1. Outcome definitions

For detailed explanation of outcome definitions, refer to outcomes section of trial registration listing: https://www.anzctr.org.au/Trial/Registration/TrialReview.aspx?id=378197&isReview=true

#### In summary

Primary endpoint*: hepatic insulin sensitivity, assessed using EGP during the low dose insulin phase of the hyperinsulinaemic-euglycaemic clamp (with deuterated glucose tracers)

*Basal Ra was previously documented as co-primary measure of hepatic insulin sensitivity. Plasma insulin concentrations during the basal phase of the clamp reflect exogenous insulin in type 1 diabetes (including infused insulin to achieve euglycaemia) and endogenous insulin in control volunteers without diabetes, who do not require infused insulin during this stage of the clamp. Thus, low-dose EGP is a more robust measure of hepatic insulin resistance in type 1 diabetes that is less sensitive to the impact of factors related to basal insulin and has been updated as the preferred hepatic insulin sensitivity outcome measure.

#### Secondary endpoints

Refer to table 1 per published protocol.^2^

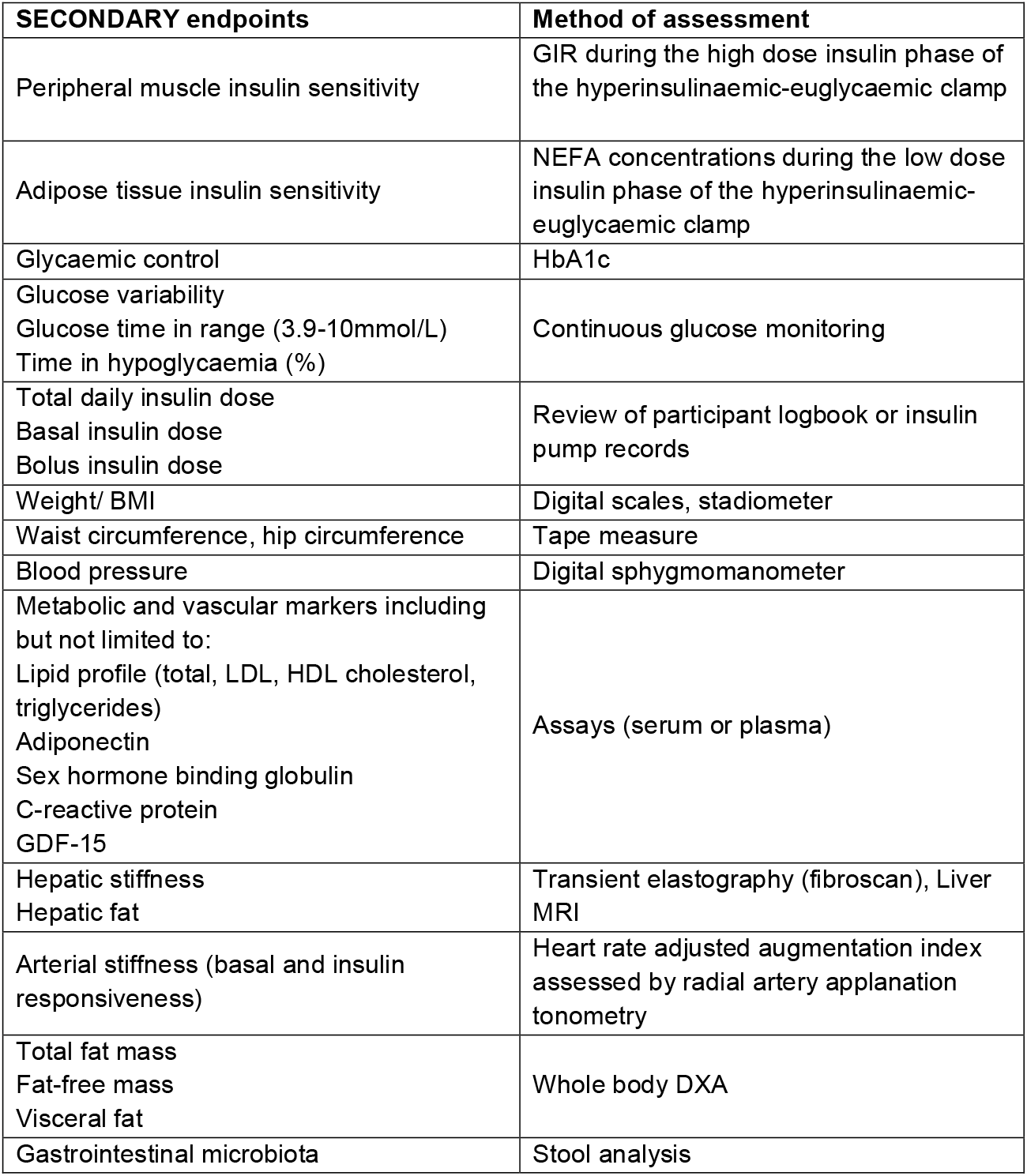
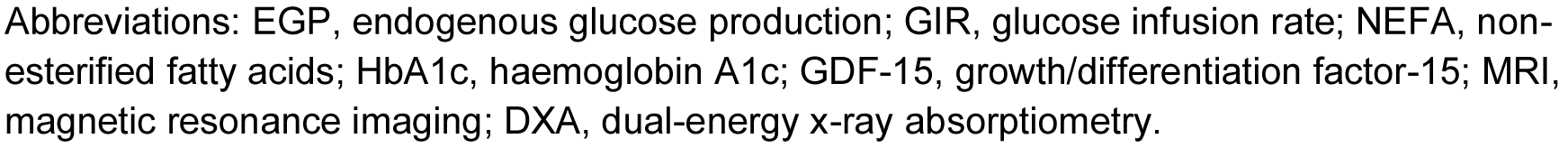

| **SECONDARY endpoints** | **Method of assessment** |
| --- | --- |
| Peripheral muscle insulin sensitivity | GIR during the high dose insulin phase of the hyperinsulinaemic-euglycaemic clamp |
| Adipose tissue insulin sensitivity | NEFA concentrations during the low dose insulin phase of the hyperinsulinaemic-euglycaemic clamp |
| Glycaemic control | HbA1c |
| Glucose variability<br>Glucose time in range (3.9-10mmol/L)<br>Time in hypoglycaemia (%) | Continuous glucose monitoring |
| Total daily insulin dose<br>Basal insulin dose<br>Bolus insulin dose | Review of participant logbook or insulin pump records |
| Weight/ BMI | Digital scales, stadiometer |
| Waist circumference, hip circumference | Tape measure |
| Blood pressure | Digital sphygmomanometer |
| Metabolic and vascular markers including but not limited to:<br>Lipid profile (total, LDL, HDL cholesterol, triglycerides)<br>Adiponectin<br>Sex hormone binding globulin<br>C-reactive protein<br>GDF-15 | Assays (serum or plasma) |
| Hepatic stiffness<br>Hepatic fat | Transient elastography (fibroscan), Liver MRI |
| Arterial stiffness (basal and insulin responsiveness) | Heart rate adjusted augmentation index assessed by radial artery applanation tonometry |
| Total fat mass<br>Fat-free mass<br>Visceral fat | Whole body DXA |
| Gastrointestinal microbiota | Stool analysis |

Abbreviations: EGP, endogenous glucose production; GIR, glucose infusion rate; NEFA, non-esterified fatty acids; HbA1c, haemoglobin A1c; GDF-15, growth/differentiation factor-15; MRI, magnetic resonance imaging; DXA, dual-energy x-ray absorptiometry.

GIR and EGP data will be adjusted to fat free mass of the individual (determined from DXA scan).

Note: Indirect calorimetry was planned for, but data collection was aborted due to infection control issues during the COVID-19 pandemic.

### 6.2. Analysis methods

Data distribution will be assessed by visual inspection of data histograms and quantile plots and supported by Shapiro-Wilk, skewness and kurtosis statistics. Normally distributed data will be presented as mean and standard deviation (SD) and analysed using parametric tests. Skewed data will be presented as median and interquartile range (IQR) then analysed with non-parametric tests or as log transformed data analysed with parametric tests. Categorical data will be presented as counts and percentages.

Effect estimates will include confidence intervals.

#### Type 1 diabetes vs control analysis

Two sample tests and Chi-square or fisher’s exact tests will be used to compare baseline characteristics between volunteers with and without type 1 diabetes. ANOVA will be used to compare characteristics between volunteers without diabetes, and type 1 diabetes defined as insulin resistant/ insulin sensitive at each tissue site. Post-hoc tests, such as Tukey’s honestly significant difference (HSD) post hoc test will be used to determine between which groups the differences occurred.

Correlations will be performed using Pearson or Spearman’s correlations according to the normality of the data, or partial correlation if controlling for variables, to determine relationships between insulin sensitivity and cardiometabolic variables.

We will use regression models to:

1. Determine predictors of insulin resistance
2. Determine if insulin resistance is a predictor of cardiometabolic outcomes (eg. hbA1c, arterial stiffness, visceral adipose tissue etc), whilst controlling for clinically relevant factors (e.g age, sex)

#### Primary outcome

The primary analysis will compare intervention groups (metformin vs placebo) to assess whether there is a difference in hepatic insulin resistance (LD-EGP) after 26 weeks. The mean change in LD-EGP (delta EGP) from baseline to 26 weeks will be assessed using a generalised linear mixed model. These models will include a random intercept for each individual, with absolute LD-EGP as the response variable at each time point. The distribution and link will be determined empirically by residual versus fixed-effect prediction plots, likelihood ratio and Akaike’s Information Criterion (AIC). Time, treatment group, and group by time interaction are pre-specified predictor variables. These same models will also be presented in a re-parameterised form with time, and group by time interaction (not including a main effect for treatment group), as group differences in the response variable at baseline can obscure the interpretation of the group by time interaction.^11^

The primary outcome for LD-EGP will be the treatment by time interaction.

We will also report the pre-post difference (with 95% confidence intervals) of the estimated marginal means with treatment group as the pairwise contrast.

We will systematically assess models as:

1. Unadjusted
2. Minimally adjusted using select physically plausible covariates (age, sex, baseline HbA1c and baseline BMI).

#### Secondary outcomes

Secondary endpoints that relate to a change from baseline measurement will be analysed as per primary endpoint analyses.

Correlations between change in insulin resistance and change in cardiometabolic factors or laboratory measurements will be analysed. We will explore the association between metabolic parameters and the response to metformin intervention.

We nominate a limited number of plausible predictor variables for adjusted analyses including:

○ Age, HbA1c, BMI, waist and hip circumference, SBP, DBP, sex
○ Triglycerides, LDL, HDL, IGF-1
○ Adiponectin
○ VAT, total body fat
○ GDF-15
○ insulin delivery method (for hypoglycaemia metrics)

Assumptions for models will be checked. Regression models will be checked by visually inspecting residual plots for evidence of heteroscedasticity and linearity. Other model diagnostics such as the variance inflation factor (VIF) for collinearity will be checked.

### 6.3. Missing data

The nature of missing data relevant to the primary endpoint will be determined to be missing at random (MAR), missing completely at random (MCAR) or missing not at random (MNAR). Multiple imputation chained equation (MICE) will be applied if it is deemed appropriate to use the missing at random assumption (MAR). If it is deemed more appropriate to use the missing completely at random assumption (MCAR), a complete case analysis approach will be used and standard mixed model analyses will deal with the missing data without imputation, with pairwise deletions where appropriate. If data is MNAR, an appropriate modelling strategy will be developed based on the estimated missingness mechanisms.

Data below the limit of quantification (LoQ) for an assay will be substituted with a constant derived by the following equation: LoQ/√2

Ie LOQ divided by the square root of 2.

Loss to follow up and volunteer drop out: the timing and reason of drop out will be recorded and presented in the CONSORT diagram within the appropriate treatment arm.

### 6.4. Additional analysis

Further exploratory analyses will be conducted if relevant.

In the event that non-essential secondary analyses are not available at the time of primary analyses, then analysis of that data will be presented separately.

### 6.5. Harms

Adverse events are assessed at regular intervals including at the time of study drug dose titration, and at 3 and 6 months. Volunteers will be asked about gastrointestinal side effects, severe hypoglycaemia, and provided opportunity to disclose other symptoms or hospital presentations. Volunteers are encouraged to contact the study team ad hoc if adverse effects arise outside these scheduled timepoints. The time of ad hoc reviews will be noted in the study file.

Gastrointestinal side effects and severe hypoglycaemia will be coded as binary events (present/ absent since starting study medication).

### 6.6. Statistical software

IBM SPSS statistical package version 28.0.1.0 will be used. RStudio may be used for select endpoint analysis if SPSS does not offer the required options for generalised linear mixed modelling appropriate to the nature of the data.

## Data Availability

Data may be made available on a case-by-case basis at the discretion of the primary sponsor and pending ethics committee approval.

## 7. OTHER

